# Functional Severity Determines Return to Sport After Ankle Sprain in Young Athletes

**DOI:** 10.64898/2026.05.03.26352308

**Authors:** Shinsuke Sakoda, Kouichi Kajiwara, Arata Yoshida, Kimiaki Kawano

**Affiliations:** Department of Sports Medicine, Ashiya Central Hospital, Fukuoka, Japan; Department of Orthopaedic Surgery, Ashiya Central Hospital, Fukuoka, Japan

**Keywords:** ankle injuries, athletes, prognosis, recovery of function, walking, return to play decision-making

## Abstract

**Objectives:** To determine whether early functional severity at presentation explains variability in return to sport (RTS) after ankle sprain in young athletes, compared with sprain subtype and injury mechanism.

**Design:** Retrospective cohort study.

**Methods:** Athletes aged ≤22 years with acute ankle sprains were identified from a prospectively maintained institutional database. Surgically treated cases were excluded. Functional severity at presentation was classified into three grades based on the ability to continue sports participation and ambulate immediately after injury. Injury mechanisms were categorized as high-energy deceleration (HED) or non-HED. RTS was analyzed as time to return and as prolonged RTS (≥4 weeks). Multivariable logistic regression was performed to identify factors independently associated with prolonged RTS.

**Results:** A total of 437 cases were included. Median RTS was 2.0 weeks (interquartile range, 0.0–4.0), and prolonged RTS occurred in 33.0% of cases. RTS duration increased stepwise with greater functional severity (p < 0.001). In multivariable analysis, functional severity was strongly associated with prolonged RTS (Grade 2: adjusted odds ratio [OR], 3.58; 95% confidence interval [CI], 2.07–6.19; Grade 3: adjusted OR, 24.53; 95% CI, 10.67–56.43; p < 0.001), and age was also independently associated (adjusted OR, 1.19 per year; 95% CI, 1.11–1.27; p < 0.001). Sprain subtype and injury mechanism were not independently associated with RTS after adjustment.

**Conclusions:** Early functional severity at presentation is the primary determinant of RTS after ankle sprain in young athletes. Apparent differences related to sprain subtype and injury mechanism are largely explained by initial functional impairment.

## Introduction

Ankle sprains are among the most common injuries in young athletes and are frequently encountered in clinical practice. Despite being often treated as a relatively uniform condition, clinical recovery after ankle sprain varies substantially, particularly in terms of return to sport (RTS). However, prognostic factors associated with recovery after ankle sprain remain inconsistent and incompletely understood across studies. ^1, 2^

Traditionally, this variability has been interpreted based on structural classifications, such as sprain subtype (lateral, medial, syndesmotic, or combined), or on injury mechanism, including high-energy deceleration (HED) movements such as cutting, landing, and rapid stopping. ^3, 4^ These factors are assumed to reflect differences in mechanical loading at the time of injury and are therefore commonly considered when estimating prognosis.

However, such classifications describe how the injury occurred or which structures are involved, rather than how severely the athlete’s functional capacity is impaired at presentation. From a clinical perspective, early functional status—such as the ability to continue sports participation or to ambulate—may provide a more direct representation of the overall impact of injury on load-bearing capacity and neuromuscular function. ^5–7^

Early functional severity at presentation can be conceptualized as an integrated clinical construct that reflects the combined effects of pain, structural injury burden, neuromuscular inhibition, and perceived instability. ^5–8^ As such, it may better capture the immediate functional consequences of injury than mechanism- or structure-based classifications. In contrast to structural severity classifications, which describe anatomical injury patterns, functional severity captures the immediate functional consequences of injury, which may not be fully explained by structural findings alone.

Importantly, apparent differences in RTS observed across sprain subtypes or injury mechanisms may not represent independent prognostic effects, but rather reflect underlying differences in early functional impairment at presentation. If so, functional severity may serve as the primary determinant of recovery, while subtype and mechanism act as indirect descriptors of injury characteristics.

Clarifying this relationship is clinically relevant, as it may inform which factors should be prioritized when estimating prognosis and guiding RTS decisions in the acute setting.

Therefore, the purpose of this study was to determine whether early functional severity at presentation explains variability in RTS after ankle sprain in young athletes, in comparison with sprain subtype and injury mechanism.

We hypothesized that functional severity would be the primary determinant of RTS, and that apparent differences associated with subtype and mechanism would be largely explained by differences in early functional impairment.

## Methods

### Study Design and Setting

This retrospective cohort study was conducted using a prospectively maintained institutional database of sports-related injuries. The database includes athletes aged ≤22 years who presented with acute lower-extremity injuries between January 2017 and November 2025 at a single sports medicine center.

### Participants

Patients diagnosed with ankle sprain based on clinical findings at initial presentation were eligible for inclusion.

To ensure a homogeneous cohort reflecting the natural clinical course of ankle sprains, patients who underwent surgical treatment—typically involving syndesmotic instability or marked talocrural joint instability requiring operative management—were excluded from the analysis.

To ensure independence of observations, only the initial injury per athlete per side was included. When multiple injuries occurred on the same side, the earliest recorded event was defined as the index injury.

Cases with missing data for injury mechanism, functional severity at presentation, or RTS were excluded from the respective analyses.

### Injury Classification

Ankle sprains were classified based on clinical diagnosis at initial presentation into the following categories: lateral ankle sprain, medial ankle sprain, high ankle sprain, and combined sprain. Classification was determined by the treating physician based on clinical findings, with imaging used when clinically indicated.

### Injury Mechanism

Injury mechanisms were classified based on clinical records into two categories:

High-energy deceleration (HED): injuries associated with rapid deceleration, cutting, or landing under weight-bearing conditions

Non-HED: all remaining mechanisms

HED was defined as a movement involving rapid deceleration under load, such as landing, cutting, or sudden stopping, irrespective of contact.

This classification was applied irrespective of contact or non-contact mechanisms, as the focus was on the presence of rapid deceleration under load rather than the mode of external force application.

### Functional Severity at Presentation

Functional severity at presentation was assessed based on the athlete’s functional status at the time of injury, including the ability to continue sports participation and to ambulate immediately after injury. This information was obtained from clinical records reflecting on-field or immediate post-injury evaluation. This classification was intended to reflect early functional impairment at the time of injury rather than structural injury severity.

Functional severity was categorized into three predefined grades:

Grade 1: able to continue sports participation

Grade 2: able to ambulate but unable to continue sports

Grade 3: unable to ambulate or bear weight

This classification captures the immediate functional consequences of injury, integrating multiple clinical components including pain, load-bearing capacity, neuromuscular function, and perceived instability.

All assessments were based on clinical records reflecting the athlete’s condition at the time of injury. Accordingly, functional severity should be interpreted as a pragmatic indicator of immediate functional impact rather than a fixed or purely objective measure.

### Return to Sport

RTS was defined as the time from injury to unrestricted return to full sports participation, as determined by the treating physician.

In clinical practice, RTS decisions were based on a stepwise progression of functional recovery, with gradual increases in loading as outlined in Table 3, and were made through agreement between the treating physician and physical therapist.

This definition reflects real-world clinical decision-making rather than a standardized performance-based assessment.

### Statistical Analysis

Descriptive statistics were calculated for baseline characteristics. Continuous variables are presented as mean ± standard deviation or median with interquartile range, as appropriate, and categorical variables as counts and percentages.

Differences in RTS across functional severity groups and sprain subtypes were assessed using appropriate statistical tests.

To identify factors associated with prolonged RTS, multivariable logistic regression analyses were performed. Separate models were constructed to evaluate the independent effects of sprain subtype and injury mechanism. Each model included age, sex, and functional severity at presentation as core covariates, with either sprain subtype or injury mechanism (HED vs non-HED) entered as an additional variable.

Adjusted odds ratios (ORs) with 95% confidence intervals (CIs) were calculated. Statistical significance was set at p < 0.05.

All analyses were performed using standard statistical software.

This study was approved by the institutional review board (approval number: 7-ACH-11-2) and was conducted in accordance with the Declaration of Helsinki. The requirement for informed consent was waived due to the retrospective nature of the study.

## Results

### Study Population

A total of 2,129 acute lower-extremity sports injuries were identified in the institutional database. Among these, 677 ankle sprain cases were identified. After excluding 18 surgically treated cases, 659 conservatively managed cases remained. Among these, 132 non-index injuries were excluded, leaving 527 index ankle sprain injuries. Of these, RTS data were available for 437 cases, which were included in the RTS analysis (Figure 1).

**Figure 1.**
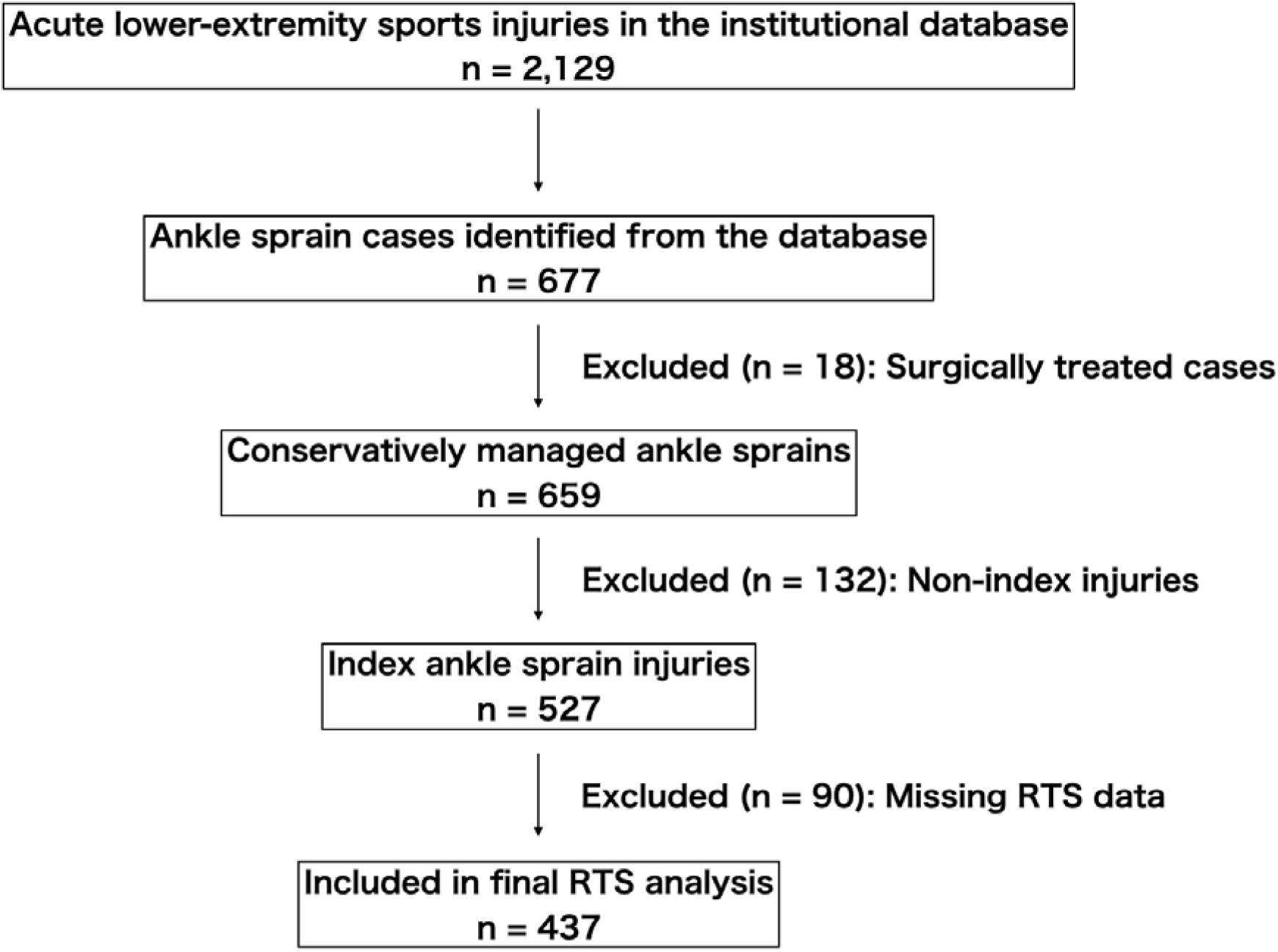
Flow diagram of patient selection and inclusion for analysis. A total of 677 ankle sprain cases were identified from the institutional database. After excluding 18 surgically treated cases, 659 conservatively managed cases remained. Among these, 132 non-index injuries were excluded to ensure independence of observations, leaving 527 index ankle sprain injuries for inclusion in the study cohort. Of these, return-to-sport (RTS) data were available for 437 cases, which were included in the RTS analysis.

The baseline characteristics of the study population are summarized in Table 1. The mean age of the analyzed cohort was 15.5 ± 3.8 years, and 331 patients (75.7%) were male.

**Table 1.** Baseline Characteristics Stratified by Prolonged RTS (≥4 weeks).

| **Variable** | **Total**<br>**(n=437)** | **RTS <4 weeks**<br>**(n=293)** | **RTS $\geq 4$ weeks**<br>**(n=144)** | **p-value** |
| --- | --- | --- | --- | --- |
| **Demographics** |  |  |  |  |
| Age, years (mean ± SD) | 15.5 ± 3.8 | 14.8 ± 3.6 | 16.8 ± 3.9 | <0.001 |
| Male, n (%) | 331 (75.7) | 219 (74.7) | 112 (77.8) | 0.51 |
| Functional severity at injury, n (%) |  |  |  | <0.001 |
| Grade 1 | 156 (35.7) | 134 (45.7) | 22 (15.3) |  |
| Grade 2 | 230 (52.6) | 148 (50.5) | 82 (56.9) |  |
| Grade 3 | 51 (11.7) | 11 (3.8) | 40 (27.8) |  |
| Injury mechanism, n (%) |  |  |  | 0.86 |
| HED | 196 (44.9) | 131 (44.7) | 65 (45.1) |  |
| Non-HED | 241 (55.1) | 162 (55.3) | 79 (54.9) |  |
| Sprain subtype, n (%) |  |  |  | <0.001 |
| Lateral | 321 (73.5) | 222 (75.8) | 99 (68.8) |  |
| Medial | 47 (10.8) | 40 (13.7) | 7 (4.9) |  |
| High ankle | 44 (10.1) | 26 (8.9) | 18 (12.5) |  |
| Combined | 25 (5.7) | 5 (1.7) | 20 (13.9) |  |

RTS = return to sport. HED = high-energy deceleration.
Continuous variables were compared using the Kruskal–Wallis test, and categorical variables were compared using the chi-square test.
Subtype-related differences should be interpreted in the context of functional severity at presentation, which represents the primary determinant of RTS.

Functional severity at presentation was classified as Grade 1 in 156 patients (35.7%), Grade 2 in 230 (52.6%), and Grade 3 in 51 (11.7%). The diagnostic distribution consisted predominantly of lateral ankle sprains (321 cases, 73.5%), followed by medial ankle sprains (47, 10.8%), high ankle sprains (44, 10.1%), and combined sprains (25, 5.7%).

### Return-to-Sport Duration According to Functional Severity at Presentation

RTS duration increased significantly with greater functional severity at presentation (p < 0.001). Median RTS was 0 weeks (interquartile range [IQR], 0–1) in Grade 1, 2 weeks (IQR, 1–5) in Grade 2, and 6 weeks (IQR, 4–8) in Grade 3 (non-ambulatory / non–weight-bearing injuries).

RTS duration according to functional severity at presentation is shown in Figure 2. The proportion of prolonged RTS (≥4 weeks) increased stepwise with severity: 14.1% in Grade 1, 35.7% in Grade 2, and 78.4% in Grade 3.

**Figure 2.**
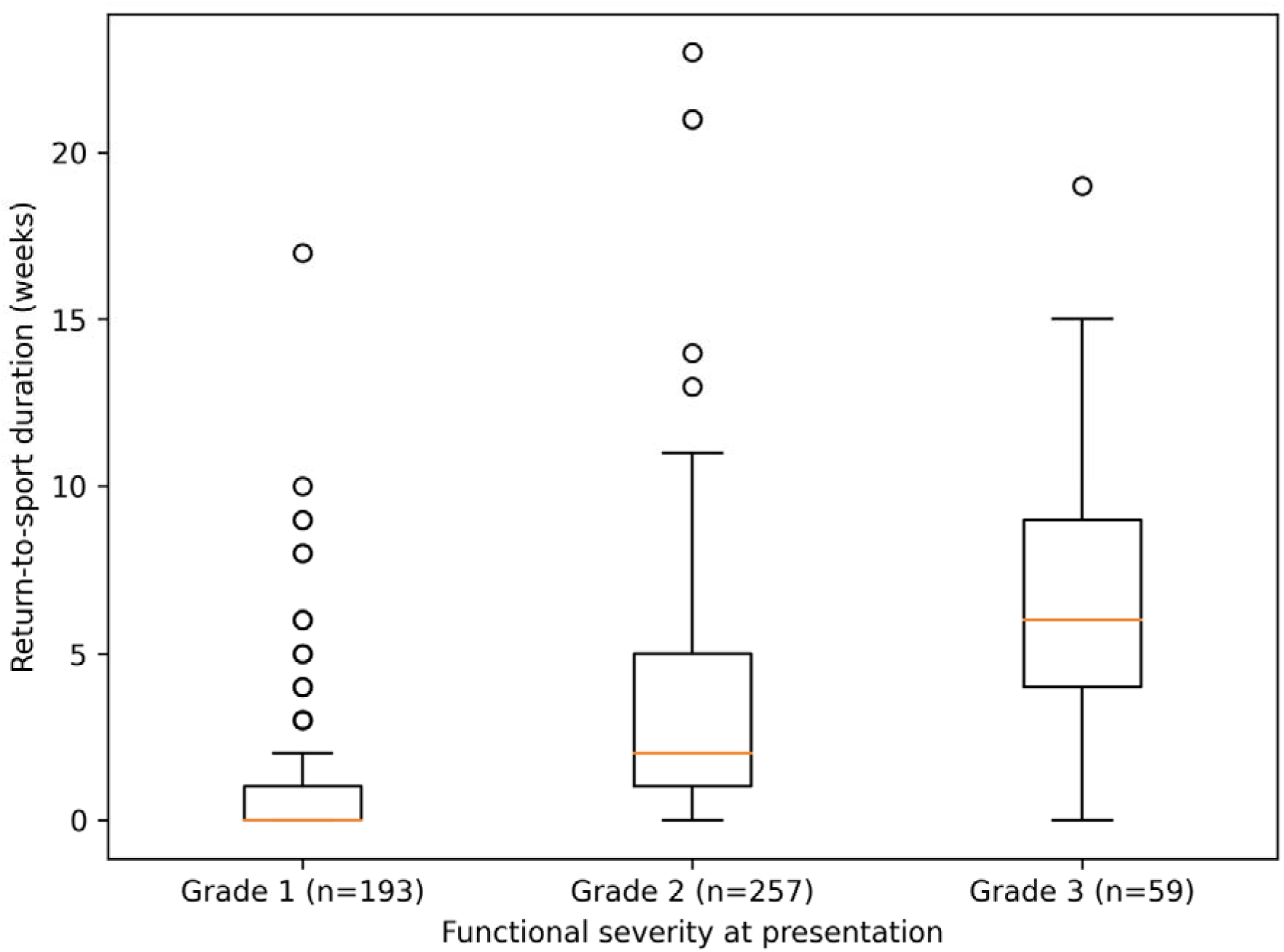
Return-to-sport (RTS) duration according to functional severity at presentation. Box plots represent the median and interquartile range, with whiskers indicating the full data range. RTS duration increased stepwise across severity grades, with Grade 1 showing the shortest recovery time and Grade 3 the longest. Grade 3 injuries represent non-ambulatory (non–weight-bearing) status at presentation. This stepwise pattern was consistent and represents the primary gradient of RTS recovery in this cohort.

This stepwise pattern was consistent across the cohort and represented the primary gradient of RTS recovery.

### Clinical Profiles and Return-to-Sport According to Sprain Subtype

Functional severity at presentation differed across sprain subtypes. Combined sprains were more frequently associated with higher severity grades, whereas medial sprains tended to present with lower functional impairment. Lateral and high ankle sprains demonstrated intermediate patterns.

RTS duration also differed significantly across sprain subtypes (p < 0.001). Combined sprains demonstrated longer recovery times, whereas medial sprains showed shorter RTS durations. Lateral and high ankle sprains showed intermediate recovery patterns.

The proportion of prolonged RTS (≥4 weeks) was highest in combined sprains (20/25, 80.0%) and lowest in medial sprains (7/47, 14.9%), with lateral (99/321, 30.8%) and high ankle sprains (18/44, 40.9%) showing intermediate proportions.

These subtype-related differences in RTS were accompanied by corresponding differences in functional severity at presentation.

RTS duration according to sprain subtype is shown in Figure 3.

**Figure 3.**
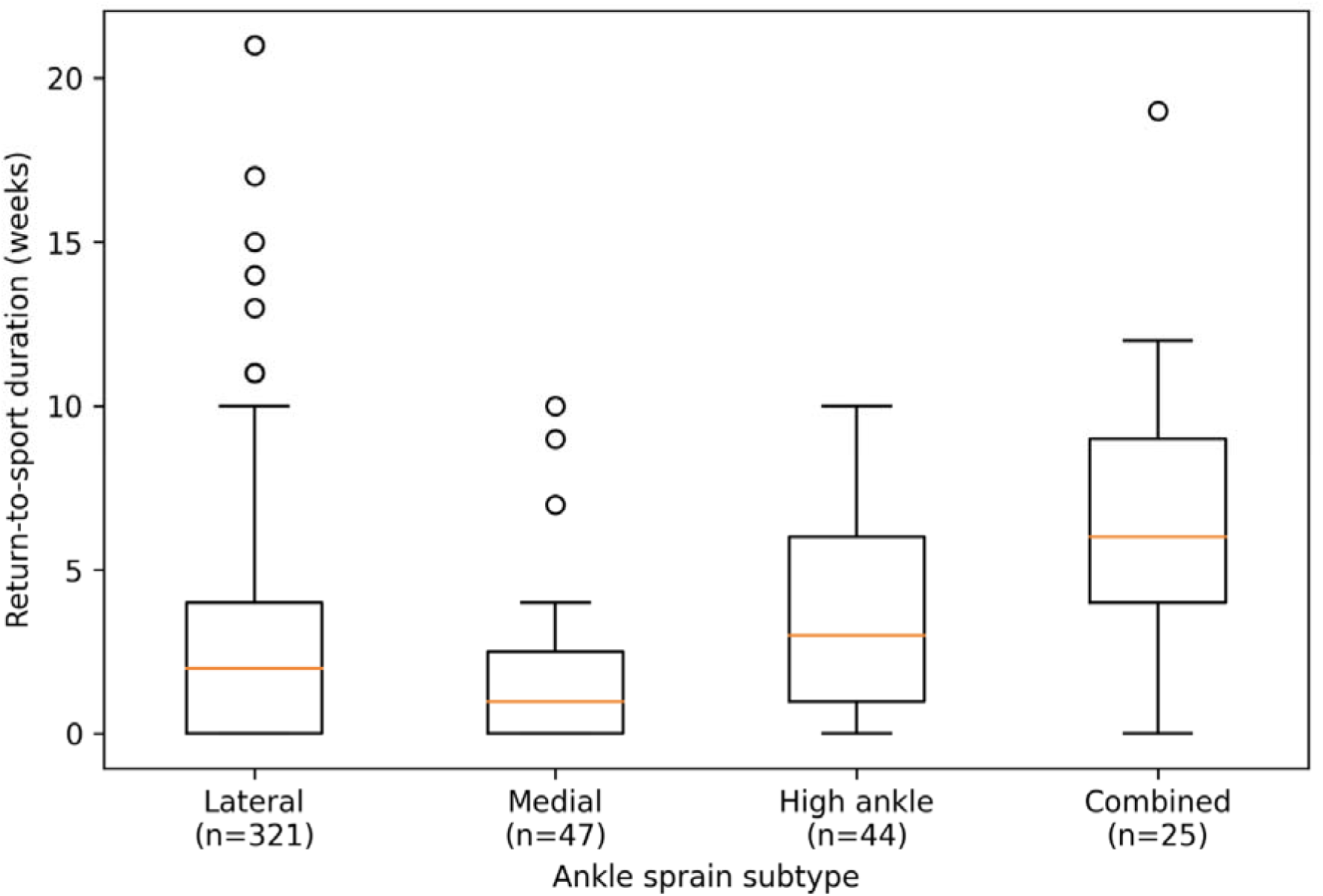
Return-to-sport (RTS) duration according to ankle sprain subtype. Box plots represent the median and interquartile range, with whiskers indicating the full data range. RTS duration differed among subtypes, with combined sprains showing longer recovery and medial sprains shorter recovery. However, these differences should be interpreted in the context of functional severity at presentation, as subtype-related differences were attenuated after adjustment for functional severity (see Table 2A).

### Factors Associated with Prolonged Return to Sport

Multivariable logistic regression analysis demonstrated that functional severity at presentation and age were independently associated with prolonged RTS.

Compared with Grade 1 injuries, Grade 2 injuries were associated with higher odds of prolonged RTS (adjusted odds ratio [OR], 3.58; 95% confidence interval [CI], 2.07–6.19; p < 0.001), and Grade 3 injuries showed a markedly stronger association (adjusted OR, 24.53; 95% CI, 10.67–56.43; p < 0.001). Age was also independently associated with prolonged RTS (adjusted OR, 1.19 per year; 95% CI, 1.11–1.27; p < 0.001).

In contrast, sprain subtype was not independently associated with RTS after adjustment for functional severity.

The results of the multivariable analysis are shown in Table 2A.

**Table 2A.** Multivariable Analysis of Factors Associated With Prolonged RTS (Including Sprain Subtype).

| **Variable** | **Adjusted OR** | **95% CI** | **p-value** |
| --- | --- | --- | --- |
| Age (per year) | 1.19 | 1.11–1.27 | <0.001 |
| Sex (male) | 1.38 | 0.77–2.47 | 0.28 |
| Functional severity |  |  | <0.001 |
| Grade 2 vs Grade 1 | 3.58 | 2.07–6.19 | <0.001 |
| Grade 3 vs Grade 1 | 24.53 | 10.67–56.43 | <0.001 |
| Sprain subtype |  |  | 0.11 (overall) |
| Medial vs Lateral | 0.40 | 0.10–1.10 | 0.07 |
| High ankle vs Lateral | 0.81 | 0.30–2.10 | 0.66 |
| Combined vs Lateral | 2.33 | 0.80–6.80 | 0.12 |

After adjustment for functional severity at presentation, sprain subtype was not independently associated with prolonged RTS.

### Injury Mechanism and Return to Sport

Injury mechanism, including HED, was not associated with prolonged RTS in multivariable analysis (adjusted OR, 0.96; 95% CI, 0.60–1.54; p = 0.85).

These findings indicate that injury mechanism did not provide additional prognostic value beyond functional severity at presentation (Table 2B).

**Table 2B.**
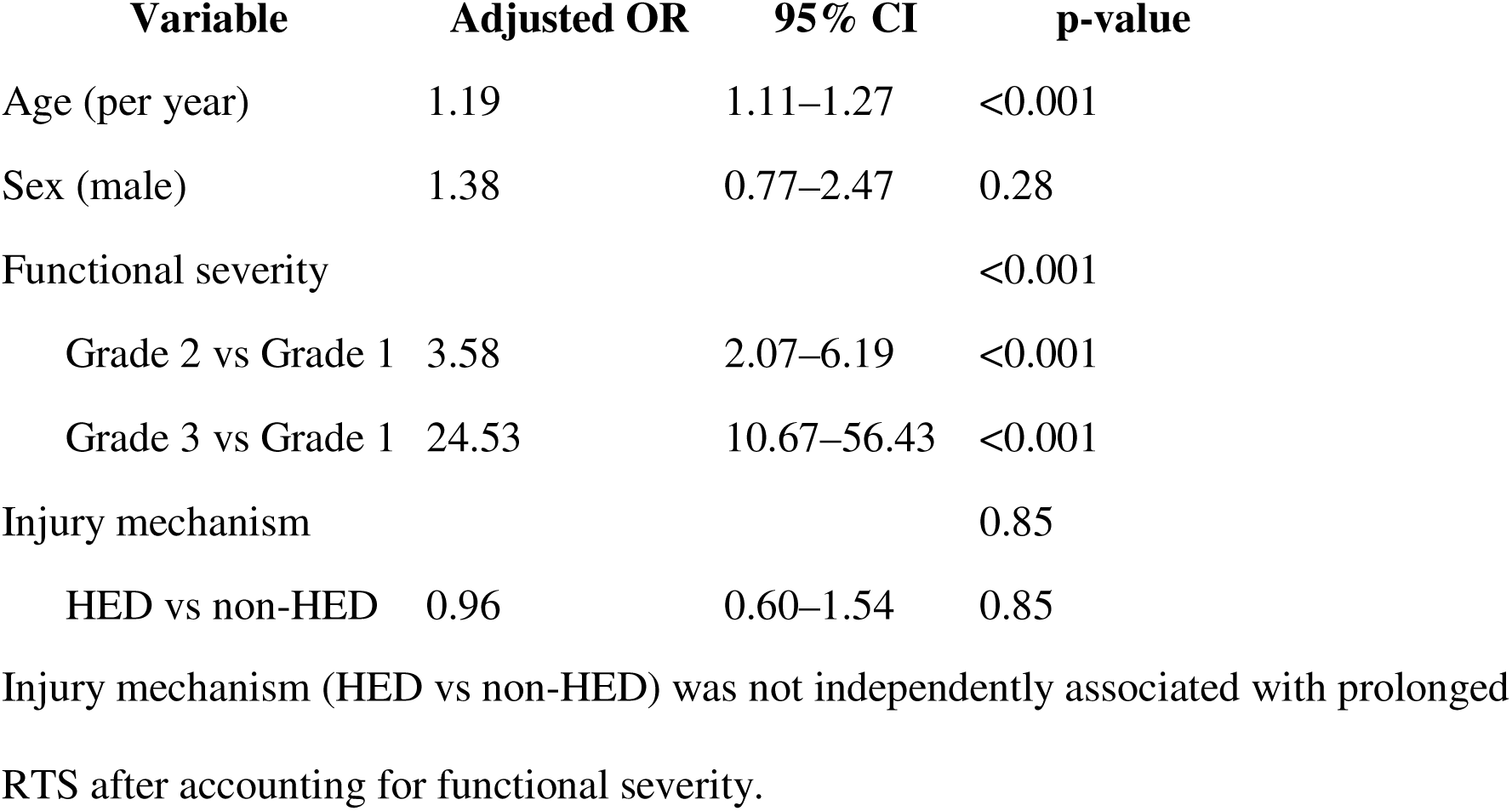
Multivariable Analysis of Factors Associated with Prolonged RTS (Including Injury Mechanism).

## Discussion

### Principal Findings

The principal finding of this study is that RTS variability after ankle sprain in young athletes was primarily determined by early functional severity at presentation. RTS duration increased in a clear stepwise manner across severity grades, and functional severity remained the strongest independent predictor of prolonged RTS after adjustment. This finding is consistent with prior studies demonstrating that early functional status, particularly weight-bearing capacity and functional limitations, provides clinically meaningful prognostic information following ankle sprain. ^6, 7, 9^ In contrast, structural classifications and injury mechanism alone appear to have limited independent prognostic value once early functional impairment is considered. ^5, 6^

Although apparent differences in RTS were observed across sprain subtypes, these differences were substantially attenuated after accounting for functional severity and were no longer statistically significant. This suggests that subtype-related differences in recovery are largely explained by differences in initial functional impairment rather than by independent effects of anatomical injury patterns.

Similarly, injury mechanism, including HED, did not provide additional prognostic value. Taken together, these findings indicate that early functional severity represents the primary determinant of short-term recovery, whereas subtype and mechanism do not independently influence RTS once functional impairment is considered.

Functional severity at presentation reflects the athlete’s immediate functional capacity in the acute phase after injury, including basic locomotion and load tolerance. In contrast, RTS is determined after progressive rehabilitation and repeated functional assessments, representing a later clinical decision regarding sport-specific readiness. ^10, 11^ Therefore, functional severity at presentation should be interpreted as an indicator of initial functional impact rather than a component or early stage of RTS.

### Conceptual Interpretation of Functional Severity

An important conceptual consideration is that the association between functional severity at presentation and RTS should not be interpreted as a tautological relationship. Rather, functional severity may reflect an early proxy of functional impairment that influences subsequent recovery trajectories.

Although imaging modalities such as MRI provide detailed information on structural injury, their relationship with functional impairment relevant to RTS remains incompletely understood. ^5^ Therefore, the present study focused on early functional status as a clinically relevant indicator of injury impact.

These constructs were not directly measured in the present study and should be interpreted as theoretical mechanisms underlying the observed associations.

### Functional Severity as the Primary Determinant of Recovery

A key observation of this study is the robust and consistent relationship between functional severity at presentation and RTS. The stepwise increase in recovery time from Grade 1 to Grade 3 suggests that early functional impairment captures the magnitude of disruption in load-bearing capacity and neuromuscular function more directly than structural or mechanistic classifications.

In particular, Grade 3 injuries—defined by inability to ambulate and bear weight—represent a threshold of functional failure at which compensatory mechanisms are insufficient to maintain basic locomotion. This threshold likely reflects a substantial loss of dynamic stability and load transmission capacity, which in turn necessitates longer recovery before safe RTS.

Functional severity can therefore be interpreted as an integrated clinical construct that summarizes the immediate consequences of injury, including pain, structural damage, neuromuscular inhibition, and perceived instability. ^8, 9^ Its strong association with RTS suggests that this integrated functional response, rather than isolated structural or mechanistic descriptors, governs the trajectory of recovery.

### Interpretation of Subtype-Related Differences

Although RTS differed across sprain subtypes in unadjusted analyses, these differences were closely aligned with variations in functional severity at presentation. Combined sprains were more frequently associated with higher severity grades, whereas medial sprains tended to present with lower functional impairment.

After adjustment for functional severity, sprain subtype was no longer associated with RTS. This finding indicates that subtype-related differences in prognosis are largely explained by differences in initial functional impairment rather than independent effects of anatomical injury patterns.

Accordingly, sprain subtype should be interpreted as a descriptor of injury characteristics that may influence initial presentation, but not as an independent determinant of recovery once the degree of functional impairment is established.

### Limited Prognostic Value of Injury Mechanism

Injury mechanism, including HED, did not demonstrate an independent association with RTS after accounting for functional severity. While HED movements are often assumed to involve greater mechanical loading at the time of injury, the present findings suggest that such external descriptors do not translate into differences in clinical recovery once the resulting functional impairment is considered.

From a clinical perspective, injury mechanism describes how the injury occurred, whereas functional severity reflects the functional consequences of that injury. The absence of an independent effect of mechanism supports the interpretation that prognosis is determined primarily by the extent of functional disruption rather than by the nature of the inciting event.

### Clinical Implications

These findings suggest that early functional assessment provides a practical approach for estimating prognosis and guiding rehabilitation and RTS decisions after ankle sprain. A function-oriented approach based on recovery of functional capacity, rather than injury subtype or mechanism, may therefore be more appropriate. ^12–14^

### Limitations

This study has several limitations. First, it was a retrospective single-center study, which may limit generalizability. Second, RTS data were not available for all cases, and analyses were restricted to patients with recorded outcomes.

Third, imaging-based severity grading was not incorporated, and therefore the relationship between structural injury severity and functional impairment could not be directly evaluated. Accordingly, the extent to which functional severity reflects underlying structural damage remains unclear.

In addition, functional severity at presentation was assessed based on the athlete’s functional status at the time of injury. Although this approach reflects real-world on-field evaluation, formal inter-rater reliability was not assessed, and variability related to assessor judgment and contextual factors (e.g., pain, competition setting) may have influenced classification. Accordingly, functional severity should be interpreted as a pragmatic indicator of immediate functional impact rather than a fixed or purely objective measure.

Future studies should incorporate prospectively standardized assessment protocols and formally evaluate inter-rater reliability, for example using agreement statistics between independent assessors, to establish the reproducibility and generalizability of this classification.

RTS in this study was determined based on clinical judgment, which may introduce variability related to individual decision-making, sport type, and competitive level. However, in practice, RTS decisions were based on stepwise functional progression with increasing loading and were made through agreement between the physician and physical therapist. This approach reflects real-world clinical practice and provides a pragmatic operational definition of RTS in routine care settings.

Finally, the function-based progression framework presented in Table 3 was not formally validated within this study and should be interpreted as a conceptual model derived from observed recovery patterns.

**Table 3.** Criteria for progression in function-based rehabilitation.

| Phase | Functional task | Criteria for progression to the next phase |
| --- | --- | --- |
| Phase 1 | Protected weight-bearing and early mobility | Able to perform double-leg squat with pain $\leq 3/10$, without worsening swelling, and with controlled movement |
| Phase 2 | Double-leg squat | Symmetrical loading, no pain increase, controlled movement |
| Phase 3 | Split squat | Stable trunk, no valgus/varus collapse, pain $\leq 3$ |
| Phase 4 | Single-leg squat | $\geq 10$ repetitions with good alignment and balance |
| Phase 5 | Jogging | $\geq 10$–15 min continuous jogging without symptom exacerbation |
| Phase 6 | Running | Able to increase speed without pain or instability |
| Phase 7 | Sprinting | Tolerates high-speed running and deceleration |
| Phase 8 | Non-contact drills | Cutting, stopping, directional change without symptoms |
| Phase 9 | RTS | RTS including full training and competition |

This progression framework is consistent with a function-oriented approach in which rehabilitation is guided by recovery of functional capacity rather than injury subtype or mechanism.
It is presented as a conceptual model derived from observed recovery patterns in this cohort and has not been formally validated. It was not directly evaluated as an outcome measure in the present study.

### Conclusion

Early functional severity at presentation is the primary determinant of RTS after ankle sprain in young athletes. Apparent differences associated with sprain subtype and injury mechanism are largely explained by differences in initial functional impairment and do not independently influence recovery once functional severity is considered.

These findings support the use of function-based assessment at initial presentation as a clinically meaningful framework for prognosis estimation and rehabilitation planning.

### Practical Implications

- Early functional assessment provides a simple and clinically useful framework for estimating recovery after ankle sprain.
- Functional severity at presentation may be a useful indicator for prognosis.
- This early functional assessment can also be used to guide rehabilitation planning and return-to-sport decision-making.

## Data Availability

All data produced in the present study are available upon reasonable request to the authors

## Acknowledgements

The authors thank the clinical staff involved in the management and data collection of the patients included in this study.

## Use of Artificial Intelligence

The authors used ChatGPT (OpenAI) for language editing and manuscript refinement. The authors reviewed and approved all content and take full responsibility for the manuscript.

